# Comparison of SARS-CoV-2 viral load in saliva samples in symptomatic and asymptomatic cases

**DOI:** 10.1101/2021.02.12.21251229

**Authors:** Anuja Bhatta, Rebecca Henkhaus, Heather L. Fehling

## Abstract

SARS-CoV-2 infections can be symptomatic as well as asymptomatic. In this study, we analyzed 460,814 saliva samples collected from July 2020 to January 2021 for a SARS-CoV-2-specific gene target using the FDA EUA test, CRL Rapid Response™, based on reverse transcription polymerase chain reaction (RT-PCR). We measured SARS-CoV-2 viral loads using cycle threshold (Ct) values. A total of 17,813 samples tested positive for COVID-19 using self-collected saliva samples. The Ct values ranged from 11 to 40, 91.3% distributed between 22 to 38 Ct. We then compared Ct values for symptomatic and asymptomatic cases for all positive saliva samples. A total of 8,706 cases were symptomatic with an average Ct value of 29.24, and 9,107 cases were asymptomatic with an average Ct value of 30.99. Hence, SARS-CoV-2 viral loads (Ct) in saliva samples for both symptomatic and asymptomatic cases are similar.

## Introduction

Coronavirus disease 2019 (COVID-19) is a global pandemic caused by a novel severe acute respiratory syndrome coronavirus 2 (SARS-CoV-2) that has surpassed 103.6 million infections and 2.24 million deaths worldwide as of February 1, 2021. Most of the COVID-19 cases are asymptomatic, while symptomatic cases range from being mild, moderate, or severe infections, while some are fatal. Studies have shown that asymptomatic COVID-19 cases are infectious (1). One of the key strategies to limit the spread of the virus is to rapidly detect and isolate both symptomatic as well as asymptomatic cases.

Many diagnostic tests for COVID-19 assays are performed using nasopharyngeal swab samples administered by healthcare professionals. Most at-home COVID-19 testing kits use self-collected anterior nasal swabs or saliva. Anterior nasal swabs are challenging to self-administer, leading to improper sample collection that adversely affects test accuracy. By comparison, saliva-based tests are more consumer-friendly because they have an easier, more comfortable collection process that produces a consistent sample every time. For our assay, we used the CRL Rapid Response™ COVID-19 test, which is a saliva-based molecular test with FDA Emergency Use Authorization (EUA) for at-home collection. This kit comes with easy-to-use self-collection instructions and has a turnaround time of 24-48 hours upon receipt to the lab. CRL Rapid Response™ is a highly sensitive method that can detect virus early in the course of infection due to its low limit of detection (250copies/mL of saliva; EUA1201219).

CRL Rapid Response™ is based on reverse transcription polymerase chain reaction (RT-PCR), which is considered the gold standard for diagnostic assays for the detection of SARS-CoV-2. Approximate viral load is measured using cycle threshold, also referred as Ct value, which represents the number of PCR cycles required for the detection of viral target during RT-PCR. The Ct value is inversely proportional to the viral load in a sample, so lower Ct value reflects higher viral loads and vice-versa. Several studies have shown that viral load (Ct) values provide valuable information to identify the state of disease condition or disease severity and can act as an important tool to monitor the disease progression (2-4).

For this study, we analyzed the presence of the SARS-CoV-2-specfic gene target, RNA dependent RNA polymerase (RdRp), using CRL Rapid Response™ in saliva samples prospectively collected from July 2020 to January 2021. We studied the distribution of viral load (Ct) in positive COVID-19 samples. Using self-reported answers from sample donors regarding symptoms and exposure to COVID-19, as well as physician follow up, we were able to compare positive samples for symptomatic versus asymptomatic cases and their corresponding average Ct values.

## Results

### Ct value distribution in saliva samples assayed for SARS-CoV-2-specific gene target

We performed the CRL Rapid Response™ COVID-19 Saliva Test on all the samples (n=460,814 total samples positive and negative) collected from July 2020 to January 2021. Using this RT-PCR assay for SARS-CoV-2, we observed a total of 17,813 COVID-19 positive samples (3.9%). We constructed a bar chart to study the distribution of viral load (Ct) in the positive population, shown in **Figure 1**. 7.2% of positive samples had Ct value below 22, 91.3% of the positive samples had Ct value between 22 to 38, and 1.5% of positive samples had Ct value above 38. Ct values less than 25 reflect a high viral load, Ct values 25-30 indicate a medium viral load, and Ct values above 30 suggest low viral loads (5). Hence, our data demonstrated the viral load in the positive population ranged from low viral load to high viral load.

**Fig 1:**
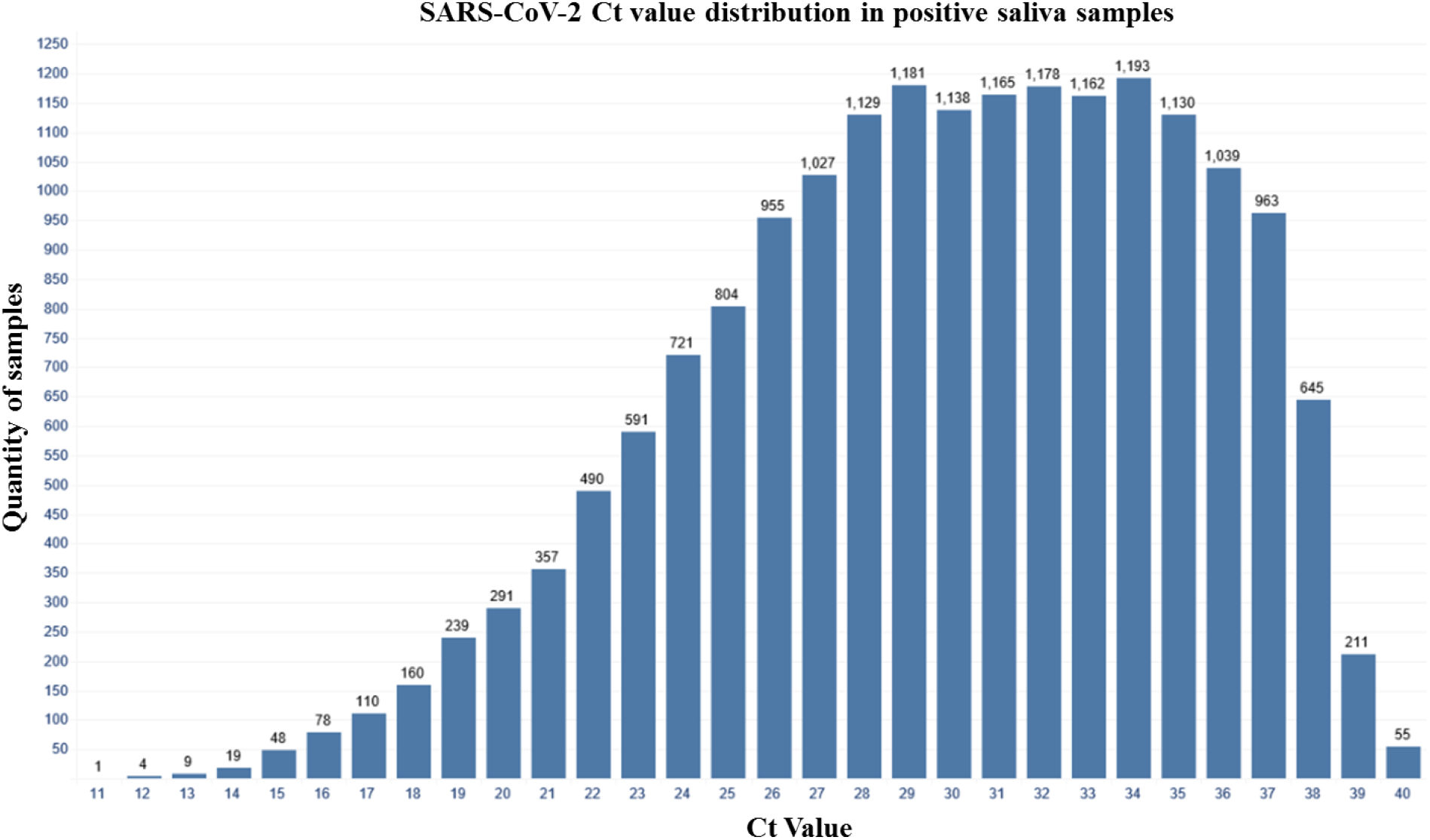
Bar chart showing Ct value distribution in saliva samples assayed for SARS-CoV-2-specific gene target. Saliva samples were analyzed using the CRL Rapid Response™ COVID-19 Saliva Test. The bar graph represents SARS-CoV-2 viral load for all COVID-19 positive samples as measured by Ct value using RT-PCR. Each bar represents the number of samples that had a Ct value in that range, for example: 12-12.99, 13-13.99, etc.

### Viral load (Ct) in saliva samples is similar in symptomatic and asymptomatic cases

Next, we performed a correlation study on our asymptomatic population to demonstrate accuracy in the testing of such cases. Asymptomatic individuals self-reported during online registration of the test that they did not meet the CDC criteria for COVID-19 symptoms or have potential exposure prior to testing, which was confirmed by post-test physician consultations. The CDC reports a wide range of symptoms for COVID-19 which includes fever or chills, cough, shortness of breathing, muscle or body aches, loss of smell/taste, sore throat and diarrhea. We determined individuals to be symptomatic if they answered ‘yes’ to any of the four questions: fever temperature greater than or equal to 100.4, persistent dry cough, experienced shortness of breath or trouble breathing and other flu like symptoms. Using these samples, we performed a correlation study as per FDA recommendations for an asymptomatic claim to be added to an emergency use authorized assay. The FDA recommends testing a minimum of 20 asymptomatic samples positive for COVID-19 and at least 100 negative samples. All specimens were required to be tested using another FDA EUA authorized molecular assay for confirmation. We included 31 asymptomatic cases positive for COVID-19 and 108 negative samples, all of which were initially tested using the CRL Rapid Response™ COVID-19 Saliva Test. Samples (n=139) were provided to P23 Labs for secondary testing since they also have FDA EUA for a saliva-based test, P23 Labs TaqPath SARS-CoV-2 Assay, leveraging the same collection device, DNA Genotek OMNIgene® ORAL (OM-505), as used by CRL Rapid Response™. P23 Labs confirmed all asymptomatic samples as positive for SARS-CoV-2 (100%) and verified all negative samples (100%) (**Table 1**). Ct values on average differed by 0.58 Ct between CRL and P23 for all COVID-19 positive asymptomatic individuals. These results demonstrate the utility of CRL Rapid Response™ COVID-19 Saliva Test in screening asymptomatic individuals given the high sensitivity and specificity of the test, as confirmed by a secondary FDA EUA test performed by an external lab.

**Table 1:**
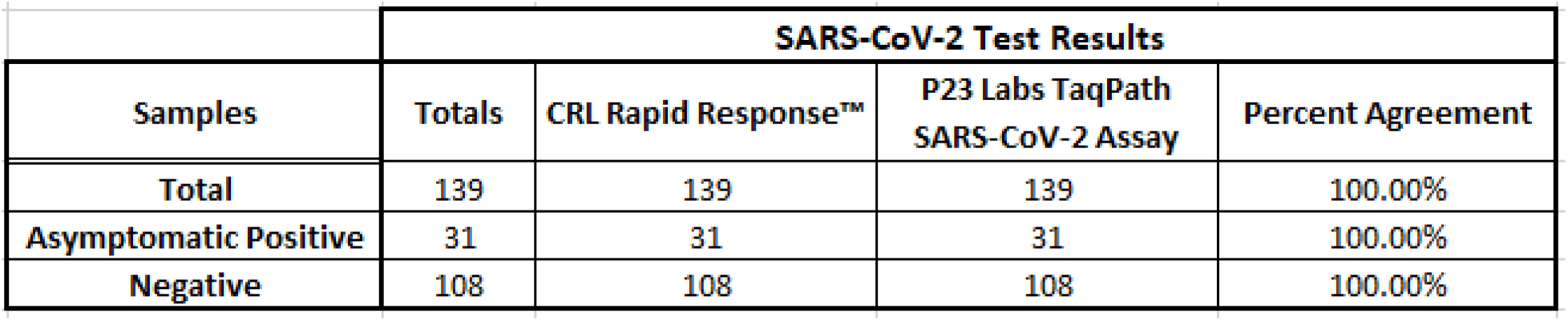
Summary of asymptomatic study data from two FDA EUA authorized saliva-based molecular assay-CRL Rapid Response™ COVID-19 Saliva test and P23 Labs TaqPath SARS-CoV-2 assay.

After confirming the accuracy of the CRL Rapid Response™ with an external lab, we investigated the relationship between viral load (Ct) and the presence or absence of symptoms in individuals testing as positive for COVID-19. Out of the 17,813 total COVID-19 positive individuals, 8,706 (48.9%) were symptomatic, and 9,107 (51.1%) were asymptomatic (**Table 2**). The symptomatic cases had an average Ct value of 29.24 and asymptomatic cases had an average Ct value of 30.99, with a difference of 1.75 Ct value between the two populations (**Table 2**). These results demonstrated that the asymptomatic population tested using CRL Rapid Response™ had no significant difference in viral load (Ct) compared to the symptomatic population.

**Table 2:** Symptomatic vs asymptomatic Ct values in saliva samples.

| Population | Average Ct | Number of Samples |
| --- | --- | --- |
| Asymptomatic | 30.99 | 9107 |
| Symptomatic | 29.24 | 8706 |
| **Difference** | 1.75 |  |

## Discussion

This study focused on analyzing the viral load (Ct) value distribution in the COVID-9 positive population using saliva as a test matrix which demonstrated similarity in Ct values in symptomatic and asymptomatic populations.

Ct values obtained from RT-PCR assays can be essential to identify COVID-19 severity and useful information to monitor the disease progression. Several studies have highlighted the correlation between Ct values and infectivity of SARS-CoV-2. Westblade et al. found that viral load correlated with mortality in hospitalized patients as well as hospitalized patients with active cancer (2). Bullard et al. showed that viral load correlated with infectivity of SARS-CoV-2 with a decrease in infectivity by 32% for every 1 unit increase in Ct value (3). Another study showed that the host immune response is dependent on viral load and duration of infection (6). All these studies were conducted using nasopharyngeal or oropharyngeal swab as test matrix.

Regarding saliva-based molecular COVID-19 tests, a study by Silva et al. illustrated that saliva is a better predictor of disease progression as compared to nasopharyngeal swabs (4). In this study, viral load in saliva correlated with increasing levels of COVID-19 disease severity while nasopharyngeal viral loads did not statistically significantly correlate to disease severity (4). Higher saliva viral loads strongly correlated with disease severity, depletion of immune cells and acted as predictor of mortality overtime (4). Similar to our findings, Silva et al. found that saliva sample collection is an easy and comfortable process compared to nasal swab collection, which can be uncomfortable leading to improper sample collection and ultimately affecting the accuracy of the assay.

We conclude that symptomatic and asymptomatic individuals have similar viral loads indicated by Ct values. A few studies have been conducted to study the correlation of Ct values with asymptomatic cases using nasopharyngeal, anterior nares or oropharyngeal swab samples. These studies have shown that there is little difference between Ct values for symptomatic and asymptomatic cases. Kissler et al. reported a mean Ct value for symptomatic individuals of 22.2 and asymptomatic individual of 22.4 (7). Gorzalski et al. compared asymptomatic cases with all COVID-19 positive cases and demonstrated that the difference in mean Ct values was 2.08, which they determined as not statistically significant. All COVID-19 individuals had an average Ct value of 27.55 while asymptomatic cases had a Ct value of 29.63 (8). Consistent with the study of other matrices, our data using saliva as test matrix also demonstrated that the two populations have little difference between Ct values with a difference of only 1.75 Ct.

Hence, molecular tests using saliva as a matrix are equally as effective for detecting viral load in symptomatic or asymptomatic donors.

## Methods

All the steps/procedures were performed as per the CRL Rapid Response™ Emergency Use Authorization issued by the FDA (EUA1201219). Briefly, the saliva samples were collected using DNA Genotek OMNIgene® ORAL device (OM-505) for the stabilization of microbial and viral RNA. For RNA extraction, we used the Zymo Quick-DNA/RNA™ Viral MagBead kit designed for high-throughput purification of viral RNA on Tecan Fluent 480/780/1080 DreamPrep Automation Workstations. For RT-PCR, we used Co-Diagnostics Inc. Logix Smart™ Coronavirus Disease 2019 (COVID-19) reagents on Bio-Rad CFX96™ Touch Real-Time PCR Detection Systems using the Bio-Rad CFX Manager 3.1 software.

## Data Availability

The data that support the findings of this study are available from the corresponding author, [Anuja Bhatta], upon reasonable request.

